# Cardiopulmonary exercise testing to evaluate post-acute sequelae of COVID-19 (“Long COVID”): a systematic review and meta-analysis

**DOI:** 10.1101/2022.06.15.22276458

**Authors:** Matthew S. Durstenfeld, Kaiwen Sun, Peggy M. Tahir, Michael J. Peluso, Steven G. Deeks, Mandar A. Aras, Donald J. Grandis, Carlin S. Long, Alexis Beatty, Priscilla Y. Hsue

## Abstract

**Importance:** Reduced exercise capacity is commonly reported among individuals with Long COVID (LC). Cardiopulmonary exercise testing (CPET) is the gold-standard to measure exercise capacity to identify causes of exertional intolerance.

**Objectives:** To estimate the effect of SARS-CoV-2 infection on exercise capacity including those with and without LC symptoms and to characterize physiologic patterns of limitations to elucidate possible mechanisms of LC.

**Data Sources:** We searched PubMed, EMBASE, and Web of Science, preprint severs, conference abstracts, and cited references in December 2021 and again in May 2022.

**Study Selection:** We included studies of adults with SARS-CoV-2 infection at least three months prior that included CPET measured peak VO_2_. 3,523 studies were screened independently by two blinded reviewers; 72 (2.2%) were selected for full-text review and 36 (1.2%) met the inclusion criteria; we identified 3 additional studies from preprint servers.

**Data Extraction and Synthesis:** Data extraction was done by two independent reviewers according to PRISMA guidelines. Data were pooled with random-effects models.

**Main Outcomes and Measures:** *A priori* primary outcomes were differences in peak VO_2_ (in ml/kg/min) among those with and without SARS-CoV-2 infection and LC.

**Results:** We identified 39 studies that performed CPET on 2,209 individuals 3-18 months after SARS-CoV-2 infection, including 944 individuals with LC symptoms and 246 SARS-CoV-2 uninfected controls. Most were case-series of individuals with LC or post-hospitalization cohorts. By meta-analysis of 9 studies including 404 infected individuals, peak VO_2_ was 7.4 ml/kg/min (95%CI 3.7 to 11.0) lower among infected versus uninfected individuals. A high degree of heterogeneity was attributable to patient and control selection, and these studies mostly included previously hospitalized, persistently symptomatic individuals. Based on meta-analysis of 9 studies with 464 individuals with LC, peak VO_2_ was 4.9 ml/kg/min (95%CI 3.4 to 6.4) lower compared to those without symptoms. Deconditioning was common, but dysfunctional breathing, chronotropic incompetence, and abnormal oxygen extraction were also described.

**Conclusions and Relevance:** These studies suggest that exercise capacity is reduced after SARS-CoV-2 infection especially among those hospitalized for acute COVID-19 and individuals with LC. Mechanisms for exertional intolerance besides deconditioning may be multifactorial or related to underlying autonomic dysfunction.

## Introduction

After SARS-CoV-2 infection, a meaningful proportion of survivors experience persistent cardiopulmonary symptoms and exercise intolerance called “Long COVID” (LC), a type of post-acute sequelae of COVID-19 (PASC). LC may occur in 3-30% of individuals after SARS-CoV-2 infection,^1-5^ including non-hospitalized and vaccinated individuals,^6,7^ and can persist for at least 12 months.^8^

Cardiopulmonary exercise testing (CPET) is the gold-standard for measuring exercise capacity and aiding in differential diagnosis of exercise limitations.^9-11^ After measuring resting cardiopulmonary parameters, participants exercise on cycle ergometer or treadmill with measurement of gas exchange and cardiopulmonary monitoring. Measuring oxygen consumption allows for objective and reproducible determination of exercise capacity (peak VO_2_), determination of anaerobic threshold, and classification of limitations. Research CPET has provided insight into persistent symptoms after SARS,^12^ dyspnea in people living with HIV,^13^ and exercise intolerance in myalgic encephalitis/chronic fatigue syndrome (ME/CFS).^14-16^ Clinically, CPET is useful diagnostically for unexplained dyspnea^9^ and prognostically in heart failure,^17^ lung disease,^9^ and in preoperative evaluations.^18^

Published case-series suggest that SARS-CoV-2 infection is associated with reduced exercise capacity.^19,20^ Whether exercise intolerance persists and is associated with LC, and the pathophysiology of exertional intolerance in LC, are uncertain. Therefore, objectives of this systematic review and meta-analysis are to address whether adults with SARS-CoV-2 infection >3 months prior^21^ have reduced exercise capacity on CPET, including those with and without LC, and to identify potential causal pathways for reduced exercise capacity after SARS-CoV-2 infection.

## Methods

A comprehensive search was planned with a research librarian (PMT) to identify all studies that used CPET to evaluate exercise capacity in adults >3 months after SARS-CoV-2 infection including case-series, cohort studies, and baseline data from interventional studies. We searched PubMed, EMBASE, and Web of Science for studies published since 2020. Unpublished abstracts from conference proceedings, indexed preprints, and references of included studies were searched. The search strategy included terms and synonyms for the following: “COVID or SARS-CoV-2” along with “cardiopulmonary exercise test”, “CPET or CPX or CPEX,” “exercise capacity,” “VO2,” and “anaerobic threshold” tailored to each search engine (Supplemental Table 1). Searches were conducted on December 20, 2021 and rerun on May 24, 2022; pre-prints were searched through June 9, 2022. Titles and abstracts and full-text studies were reviewed for inclusion by two independent reviewers (MSD & KS); there were no disagreements after consensus discussions. Data extraction was performed independently, in duplicate, using REDCap.

**Table 1.** Studies that include Cardiopulmonary Exercise Testing > 3months after SARS-CoV-2 infection

| *First Author, Year, Country* | *Study Design/Sampling/Recruitment* | *SARS-CoV-2 Infected, n* | *Hosp., n (%)* | *LC, n (%)* | *Time since infection, days<sup>a</sup>* | *Primary Analytic Comparison* |
| --- | --- | --- | --- | --- | --- | --- |
| *Abdallah/Schaeffer, 2021, Canada<sup>63,73</sup>* | Prospective cohort | 63 | 25 (40%) | 34/49 (69%) | 125 ± 16 | Hospitalized vs. Non-hospitalized<sup>63</sup><br>Fatigue vs no fatigue<sup>73</sup> |
| *Alba, 2021, United States<sup>26</sup>* | Retrospective cohort referred for CPET from PASC Clinic | 18 | 3 (17%) | 18 (100%) | 257.5 (149-322) | PASC vs controls |
| *Ambrosino, 2022, Italy<sup>67</sup>* | Pulmonary rehab after severe COVID-19 | 36 | 36 (100%) | 36 (100%) | NR | Normal vs Reduced Exercise Capacity |
| *Aparisi, 2021, Spain<sup>53</sup>* | Prospective cohort post-hospitalization | 70 | 70 (100%) | 41 (59%) | 181 ± 42 | Persistent Dyspnea vs no residual dyspnea |
| *Barbagelata, 2021, Argentina<sup>45</sup>* | Retrospective EHR review of individuals referred for clinical CPET after COVID-19 | 200 | 39 (20%) | 112 (56%) | 80 ± 21 | LC vs no LC |
| *Blokland, 2020, Netherlands<sup>74</sup>* | Retrospective cohort of individuals in rehabilitation after mechanical ventilation | 23 | 21 (100%) | NR | NR | Descriptive |
| *Blumberg, 2022, Israel<sup>43</sup>* | Cross-sectional study | 43 | NR | NR | 119 ± 24 | Vaccinated vs unvaccinated |
| *Borrego Rodriguez, 2021, Spain<sup>46</sup>* | Non-hospitalized health care workers | 57 | 0 | 57 (100%) | >90 | Peak VO<sub>2</sub> >100% predicted vs <100% predicted |
| *Brown, 2022, United Kingdom<sup>33</sup>* | Prospective cohort of hospitalized without ICU stay, myocardial injury, or comorbidities | 40 | 40 (100%) | 20 (50%) | 106 | Self-reported normal exercise capacity vs reduced exercise capacity vs controls |
| *Cassar/Raman, 2021, United Kingdom<sup>29,35</sup>* | Prospective cohort after COVID hospitalization | 42 | 42 (100%) | NR (89% of overall study) | 180 (180-204) | Change in CPET from 2-3 months to 6 months and vs controls |
| *Clavario, 2021, Italy<sup>59</sup>* | Prospective cohort after COVID hospitalization | 200 | 200 (100%) | 160 (80%) | 107 (83.0-189.0) | Normal vs Reduced Exercise Capacity |
| *de Boer, 2021, United States<sup>60</sup>* | Retrospective case series of clinically referred CPETs | 50 | 5 (10%) | 50 (100%) | 180 ± 120 | Fatty Acid Oxidation & Lactate Production in PASC vs published cohorts |
| *Debeaumont, 2021, France<sup>40</sup>* | Retrospective case series of hospitalized COVID patients referred for CPET | 23 | 23 (100%) | 23 (100%) | 180 | Ward vs ICU |
| *Dorelli, 2021, Italy<sup>54,55</sup>* | Prospective cohort post-hospitalization without comorbidities | 28 | 28 (100%) | NR | 169 ± 28 | Exercise Ventilatory Inefficiency vs Efficiency |
| *Durstenfeld, 2022, United States<sup>25</sup>* | Prospective cohort without cardiovascular disease | 39 | 7 (18%) | 23 (59%) | 525 (465-552) | Cardiopulmonary PASC vs no Cardiopulmonary PASC |
| *Evers, 2022, Germany<sup>61</sup>* | Retrospective case series of patients referred for post-COVID exercise limitation or dyspnea | 30 | 21 (70%) | 30 (100%) | 129 | Change from CPET 1 to CPET 2 |
| *Frésard, 2022, Switzerland<sup>56</sup>* | Retrospective cohort of clinical CPETs referred for LC and persistent dyspnea | 51 | 36 (71%) | 51 (100%) | 119±89 | Dysfunctional breathing vs normal breathing |
| *Godinho, 2021, United Kingdom<sup>47</sup>* | Case series of hospitalized patients with persistent exercise limitations | 9 | 9 (100%) | 9 (100%) | Range 180-360 | Descriptive |
| *Jahn, 2021, Switzerland<sup>48</sup>* | Case series of patients with severe COVID pneumonitis attending post-hospitalization pulmonary rehab | 35 | 35 (100%) | NR | 90 | Impaired vs Normal peak VO<sub>2</sub> |
| *Johnsen, 2021, Denmark<sup>49</sup>* | Case-series of post COVID clinic referrals for CPET to evaluate symptoms | 31 | NR (60% of overall study) | NR (67% of overall study) | 90 | Non-hospitalized vs hospitalized |
| *Kersten, 2021, Germany<sup>66</sup>* | Case-series of post COVID clinic referrals for CPET if initial testing not revealing | 36 | NR (8% of overall study) | NR | 121 ± 77 | Descriptive |
| *Ladlow, 2022, United Kingdom<sup>31</sup>* | Prospective cohort of active military personnel | 113 | 35 (40%) | 61 (54%) | 159 ± 7 | Comparisons by hospitalization and persistent symptoms compared to controls |
| *Liu, 2021, China<sup>65</sup>* | Prospective post-hospitalization cohort | 41 | 41 (100%) | NR | 219 ± 11 | Pulmonary fibrosis vs no fibrosis |
| *Mancini, 2021, United States<sup>52</sup>* | Case-series of LC clinic referrals for CPET for symptoms | 41 | 9 (22%) | 41 (100%) | 267 ± 99 | Descriptive |
| *Margalit, 2022, Israel* | Nested case-control study within COVID recovery cohort | 141 | 14 (14%) | 66 (47%) | 240 ± 75 | Fatigue vs no significant fatigue |
| *Mohr, 2021, Germany<sup>58</sup>* | Case-series of post COVID clinic referrals for CPET for dyspnea | 10 | 6 (60%) | 10 (100%) | 115 | Descriptive |
| *Motiejunaite, 2021, France<sup>50</sup>* | Prospective cohort | 114 | 104 (91%) | 58 (51%) | 90 (71-106) | D<sub>LCO</sub> >75 vs ≤75 |
| *Moulson, 2022, United States<sup>32</sup>* | Case-series of young athletes referred for cardiopulmonary symptoms after COVID | 21 | NR | 21 (100%) | 90 ± 63 | Young symptomatic athletes vs historical controls |
| *Parkes, 2021, United Kingdom<sup>75</sup>* | Retrospective cohort of clinical CPETs | 12 | 9 (75%) | 12 (100%) | 182 ± 111 | Descriptive |
| *Pleguezuelo, 2021, Spain<sup>28</sup>* | Survivors of ARDS from bilateral COVID pneumonia requiring mechanical ventilation and tracheostomy | 15 | 15 (100%) | NR | 56 | Mechanical efficiency, peak VO<sub>2</sub> and power output COVID vs 3 control groups |
| *Ribeiro Baptista, 2022, France<sup>39</sup>* | Prospective cohort of severe COVID requiring hospitalization >7 days and oxygen | 105 | 105 (100%) | NR | 90 days after discharge | Normal vs Reduced Exercise Capacity |
| *Rinaldo, 2021, Italy<sup>38,51</sup>* | Prospective cohort post-hospitalization | 75 | 75 (100%) | 39 (52%) | 97 ± 26 | Normal vs Reduced Exercise Capacity |
| *Romero-Ortuno, 2022, Ireland<sup>41</sup>* | Cross-sectional study of symptomatic individuals within a prospective cohort | 80 | 14 (18%) | 80 (100%) | 320 (range 39-655) | Attaining >85% of predicted maximum HR vs not attaining >85% of predicted maximum HR |
| *Singh, 2021, United States<sup>27</sup>* | Prospective Cohort of Patients Referred for CPET from LC Clinic for unexplained exercise intolerance with negative initial workup | 10 | 1 (10%) | 10 (100%) | 330 ± 30 | LC vs controls |
| *Skjærten, 2021, Norway<sup>42</sup>* | Multicenter prospective cohort post-hospitalization | 156 | 156 (100%) | 59 (38%) | 104 (90-139) | COVID vs reference population norms and no dyspnea (mMRC=0) vs dyspnea (mMRC 1-4) |
| *Szekely, 2021, Israel<sup>30</sup>* | Prospective cohort of individuals evaluated at the emergency room for acute COVID-19 | 71 | NR | 58 (67%) | 91 ± 26 | COVID-19 vs control; asymptomatic vs symptomatic; severity of acute illness |
| *Vannini, 2021, Spain<sup>62</sup>* | Prospective cohort post-hospitalization | 41 | 41 (100%) | 29 (70%) | 180 | Severity of acute illness & Peak VO<sub>2</sub><80% vs >80% |
| *von Gruenewald et al, 2022, Sweden<sup>57</sup>* | Retrospective cohort of clinical CPETs | 20 | 8 (40%) | 20 (100%) | 217 (133-329) | Normal vs abnormal breathing pattern |
| *Vonbank, 2021, Austria<sup>36</sup>* | Prospective cohort | 100 | 18 (18%) | NR | 112 | Severity of acute infection |

Table legend: <sup>a</sup>Time since infection was typically defined as the time from symptom onset or positive PCR, but some studies used the date of admission or date of hospital discharge, and we have reported it as presented by each study either as mean±SD, median (IQR), or the mean or median if only one number is reported. Pleguezuelos et al reported time since hospital discharge and mean hospitalization of 23 days, but not time from infection to hospitalization; therefore, it was unclear whether participants were >3 months since infection. We identified an additional study by Ladlow et al<sup>64</sup> but it seemed likely that there were overlapping patients so we only included the main study.

We included studies with CPET measurement of peak VO_2_ (in ml/kg/min) in participants with SARS-CoV-2 infection at least 3 months prior. Studies were excluded if they were conducted <3 months after infection, estimated VO_2_, or only included children.

Quality was assessed independently by two reviewers (MSD & KS) using Cochrane’s Quality in Prognostic Studies tool to assess study populations (inclusion criteria and control groups), measurement quality (CPET exercise protocols, peak VO_2_ assessment, assessment of sub-maximal tests), confounding, and statistical analysis and reporting.

Random-effects meta-analysis was performed to estimate mean difference in peak VO_2_ between those with and without SARS-CoV-2 infection and with and without LC as defined by each study. Heterogeneity was assessed by examining forest plots, funnel plots, and I^2^. For studies that reported median and interquartile range (IQR), the median was taken as the mean, and standard deviation was estimated as IQR/1.35; if only subgroups were reported they were combined in accordance with the Cochrane Handbook. Because of the small number of studies, tests for publication bias were not performed.

This review was conducted according to PRISMA^22^ guidelines and was registered prospectively on PROSPERO [PROSPERO 2021 CRD42021299842] prior to beginning the search. The protocol is available at https://www.crd.york.ac.uk/prospero/display_record.php?ID=CRD42021299842.

Meta-analyses were performed using STATA version 17.1.

## Results

From our search strategy and screening (Figure 1), we identified 39 observational studies that met our inclusion criteria (Table 1) including 33 published manuscripts, 3 conference abstracts, and 3 pre-prints. We identified one study of cardiac rehabillitation^23^ with baseline CPETs reported in an included study.^24^ Most studies (33, 85%) were single-center case-series of patients attending LC clinics or cross-sectional assessments within COVID-19 recovery cohorts. Three included longitudinal CPET. Most studies performed CPET 3-6 months after infection; only one study investigated individuals > 1 year after infection.^25^ Nearly half of the studies (16/39, 41%) only included hospitalized individuals (median 73% hospitalized, range 0-100%), and most predominantly included symptomatic individuals (median 89% symptomatic, range 38-100%).

**Figure 1.**
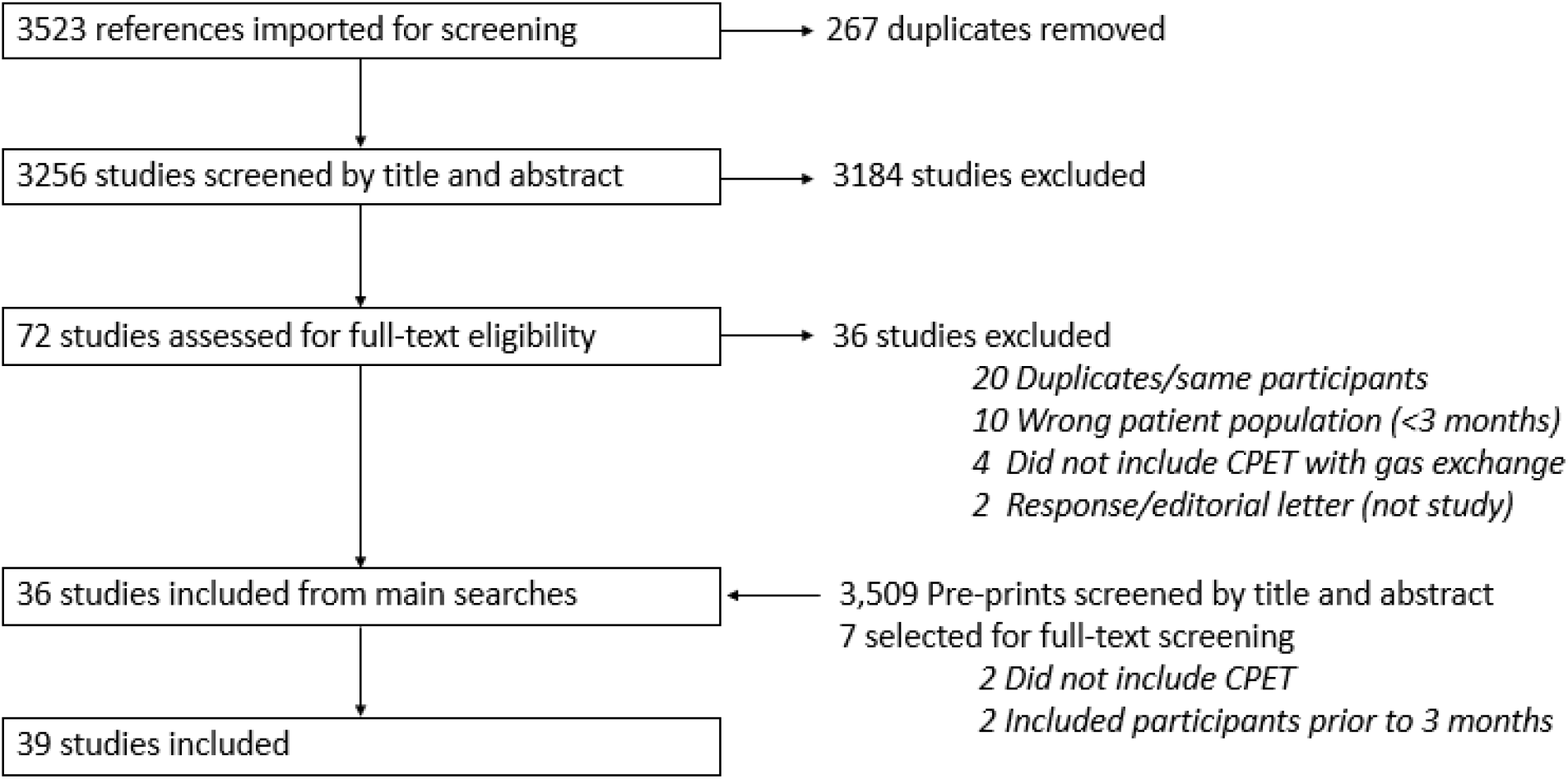
PRISMA Study Screening Diagram.

### Exercise Capacity in SARS-CoV-2 Infected Individuals Compared to Uninfected Controls

Nine studies compared 404 participants with SARS-CoV-2 to 246 uninfected controls (Table 2a). By meta-analysis (Figure 2a), SARS-CoV-2 infection was associated with 7.4 ml/kg/min lower peak VO_2_ (95%CI -11.0 to -3.7) compared to uninfected controls. The very high heterogeneity suggests that the observed variability may be due to true differences.

**Table 2.** Comparison of Peak VO_2_ between SARS-CoV-2 infected and uninfected

| *First Author, Year, Country* | *Study Design/Sampling/Recruitment* | *SARS-CoV-2 Infected, n* | *Hosp., n (%)* | *LC, n (%)* | *Time, days* | *Peak VO<sub>2</sub> after COVID-19* | *COVID-negative controls* | *Peak VO<sub>2</sub> Controls* |
| --- | --- | --- | --- | --- | --- | --- | --- | --- |
| *Alba, 2021, United States<sup>26</sup>* | Retrospective Cohort of Patients Referred for CPET from LC Clinic | 18 | 3 (17%) | 18 (100%) | 257.5 (149-322) | 20.0 (16-27)<br>85.5% (69-100) | 18 with unexplained dyspnea (historical) | 19.5 (16-23.5)<br>85% (68-100) |
| *Brown, 2022, United Kingdom<sup>33</sup>* | Prospective cohort of hospitalized without ICU stay, myocardial injury, or comorbidities | 40 | 40 (100%) | 20 (50%) | 106 | Asymp: 19.1 (15.4-23.7)<br>LC: 14.9 (13.1-16.2) | 20 (mixed historical/contemp.) | 22.3 (16.9-27.6) |
| *Cassar/Raman, 2021, United Kingdom<sup>29,35</sup>* | Prospective cohort after COVID-19 hospitalization | 42 | 42 (100%) | NR (89% overall study) | 180 (180-204) | 20.5 (17.5-26.1)<br>93.3 ± 29% | 30 (contemp.) | 28.1 (22.1-34)<br>113 ± 27% |
| *Ladlow, 2022, United Kingdom<sup>31</sup>* | Prospective cohort of active military personnel | 87 | 35 (40%) | 61 (70%) | 159 ± 72 | Hosp. LC: 29.9 ± 5.0<br>Com. LC.: 34.4 ± 7.2<br>Hosp. recovered: 32.6 ± 6.6<br>Com. recovered: 44.3 ± 7.4 | 26 (contemp.) | 43.9 ± 3.1<br>133 ± 25% |
| *Moulson, 2022, United States* | Case-series of athletes referred for LC | 21 | NR | 21 (100%) | 90 ± 63 | 44.6 ± 9.1<br>110 ± 30% | 42 (historical) | 46.4 ± 9.6<br>114 ± 23% |
| *Pleguezuelos, 2021, Spain<sup>28</sup>* | Survivors of ARDS from bilateral COVID pneumonia requiring mechanical ventilation and tracheostomy | 15 | 15 (100%), all ICU | NR | 56 | 17.3 (14.8-19.8) | 45 (15 healthy, 15 ischemic heart disease, 15 COPD) | 32.3 (28.3-36.3)<br>among healthy controls |
| *Singh, 2021, United States<sup>27</sup>* | Prospective Cohort of Patients Referred for CPET from LC Clinic for exercise intolerance with negative initial workup | 10 | 1 (10%) | 10 (100%) | 330 ± 30 | 16.7 ± 4.2<br>70 ± 11% | 10 with unexplained dyspnea & normal exercise capacity (historical) | 33.5 ± 12.9 ml/min/kg<br>131 ± 45% |
| *Szekely, 2021, Israel<sup>30</sup>* | Prospective cohort of individuals | 71 | NR<sup>a</sup> | 48 (67%) | 91 ± 26 | 1.6 ± 0.5 L/min (19.5 ml/kg/min) | 35 group matched with normal | 2.24 ± 0.9 L/min (28.1 ml/kg/min) |

|  | evaluated at the<br>emergency room<br>for acute COVID-19 |  |  |  |  |  | VO<sub>2</sub><br>(historical) |  |
| --- | --- | --- | --- | --- | --- | --- | --- | --- |
| *Vonbank,*<br>*2021,*<br>*Austria*<sup>36</sup> | Prospective<br>cohort | 100 | 18<br>(18%) | NR | 112 | Mild Acute:<br>28.2 ± 9.0<br>100.4 ±<br>24.8%<br>Severe Acute:<br>21.3 ± 6.4<br>86.1 ± 20.6% | 50<br>(historical) | 29.6 ± 7.5<br>107.7 ±<br>16.0% |

Peak VO<sub>2</sub> is reported in ml/kg/min and percent predicted when available unless otherwise noted (Szekely) as mean ± standard deviation or median (IQR) as reported by the authors. Symptoms at time of CPET (suggestive of LC) are as defined by the authors. <sup>a</sup>All patients were evaluated in the emergency department but the number or percent admitted was not reported. LC=Long COVID. NR=not reported. Hosp=Hospitalized for acute infection, Com=community (not-hospitalized) for acute infection. Contemp=Contemporary

**Figure 2.**
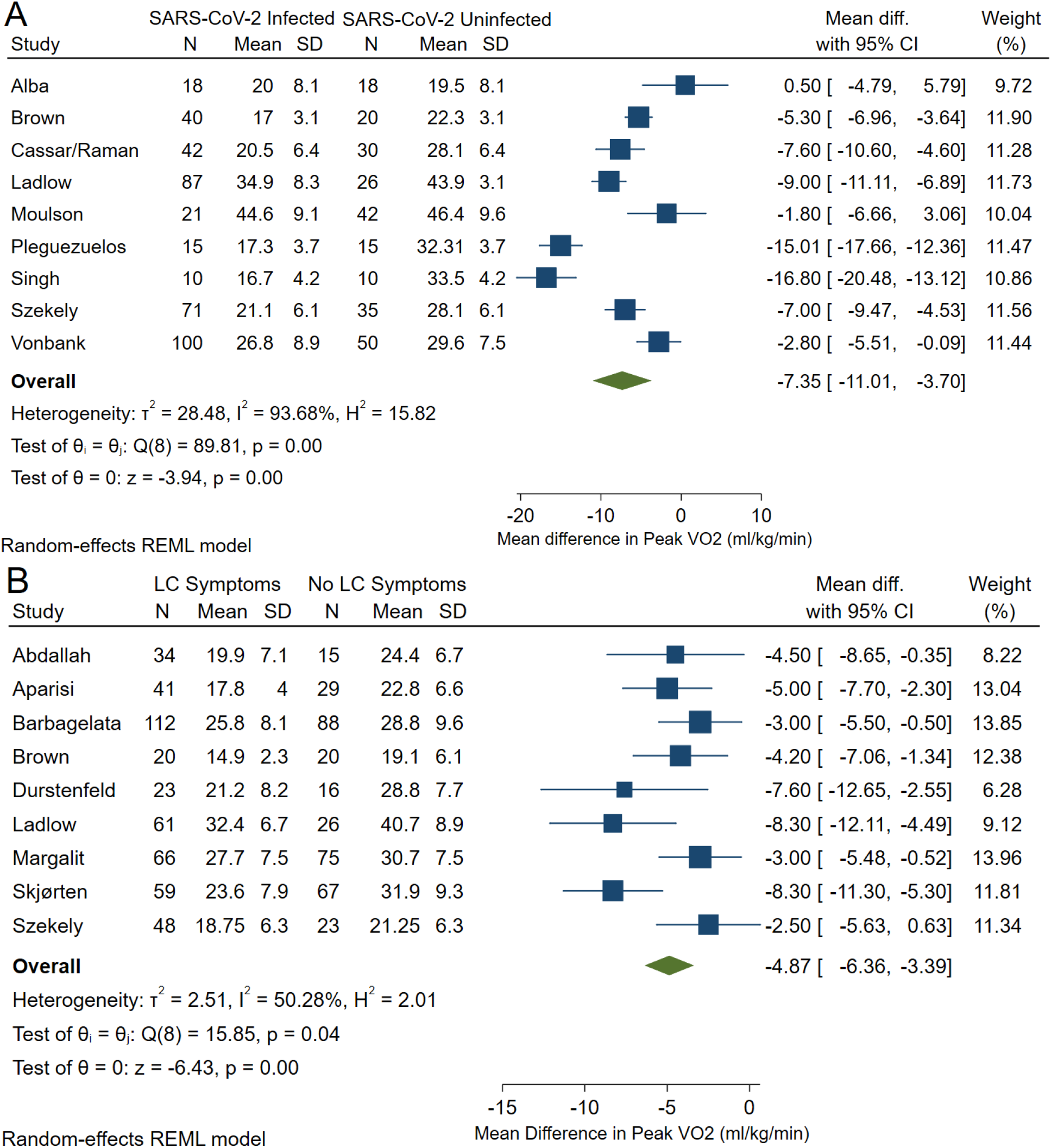
Forest plot of meta-analyses of (A) COVID vs SARS-CoV-2 uninfected controls and (B) LC Symptoms vs No LC Symptoms. Including the 9 studies of 404 individuals with SARS-CoV-2 infection and 246 individuals without SARS-CoV-2 infection, the mean difference in peak VO_2_ was -7.4 ml/kg/min (95%CI -11.0 to -3.7) by random effects meta-analysis. There was a high degree of heterogeneity with an I^2^ of 94%. Including the 9 studies of 464 individuals with LC symptoms and 359 individuals without LC symptoms (as defined by each study), the mean difference in peak VO_2_ was -4.9 ml/kg/min (95%CI -6.4 to -3.4) by random effects meta-analysis.

Heterogeneity may arise from differences in study design, participant selection, and control selection. First, case-series and case-control studies include selected portions of the spectrum of SARS-CoV-2 recovery. As case-series, Alba and Singh recruited symptomatic individuals from LC clinics^26,27^ and Pleguezuelos recruited patients after prolonged mechanical ventilation.^28^ In contrast, Cassar^29^ and Szekely^30^ enrolled cohorts during acute infection, limiting selection bias to initial severity and participant retention.

Most included participants were hospitalized for severe acute infection and most remained symptomatic, so these studies largely estimate exercise capacity among those with post-hospitalization LC. Exceptions include Ladlow and Mouslon who recruited active military personnel^31^ and athletes,^32^ respectively.

Three studies (Brown, Ladlow, Cassar) included frequency-matched contemporary controls.^31,33-35^ Five studies included age- and sex-matched historical controls with unexplained dyspnea and normal exercise capacity^27,30^ or irrespective of exercise capacity.^26,32,36^ Alba and Singh’s discrepant results may be explained by using control groups with reduced (Alba) versus known normal exercise capacity (Singh).^27^ Pleguezuelos included healthy controls, controls with ischemic heart disease, and controls with COPD.^28^

### Exercise Capacity in LC Compared to Recovered Individuals

Eighteen studies reported peak VO_2_ among individuals with LC (Supplemental Table 2) but only nine studies included those without LC (Table 3). Because the definitions of LC/PASC have evolved throughout the pandemic, the studies comparing those with and without LC used different definitions, mostly based on prevalent symptoms at CPET (Table 3).

**Table 3:**
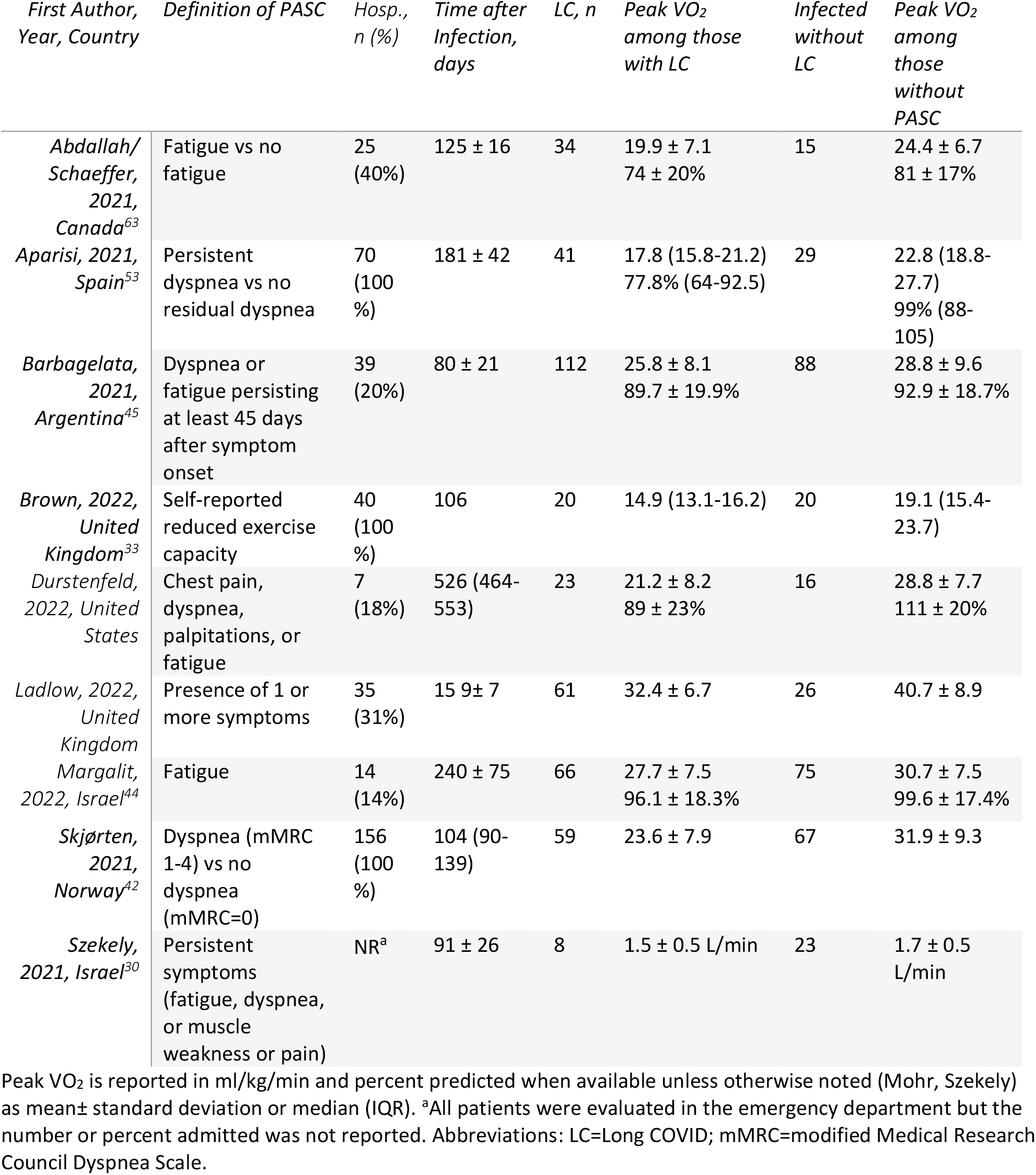
Studies with comparisons between PASC and no PASC among studies reporting VO_2_ among individuals with PASC

| *First Author, Year, Country* | *Definition of PASC* | *Hosp., n (%)* | *Time after Infection, days* | *LC, n* | *Peak VO<sub>2</sub> among those with LC* | *Infected without LC* | *Peak VO<sub>2</sub> among those without PASC* |
| --- | --- | --- | --- | --- | --- | --- | --- |
| *Abdallah/Schaeffer, 2021, Canada<sup>63</sup>* | Fatigue vs no fatigue | 25 (40%) | 125 ± 16 | 34 | 19.9 ± 7.1<br>74 ± 20% | 15 | 24.4 ± 6.7<br>81 ± 17% |
| *Aparisi, 2021, Spain<sup>53</sup>* | Persistent dyspnea vs no residual dyspnea | 70 (100 %) | 181 ± 42 | 41 | 17.8 (15.8-21.2)<br>77.8% (64-92.5) | 29 | 22.8 (18.8-27.7)<br>99% (88-105) |
| *Barbagelata, 2021, Argentina<sup>45</sup>* | Dyspnea or fatigue persisting at least 45 days after symptom onset | 39 (20%) | 80 ± 21 | 112 | 25.8 ± 8.1<br>89.7 ± 19.9% | 88 | 28.8 ± 9.6<br>92.9 ± 18.7% |
| *Brown, 2022, United Kingdom<sup>33</sup>* | Self-reported reduced exercise capacity | 40 (100 %) | 106 | 20 | 14.9 (13.1-16.2) | 20 | 19.1 (15.4-23.7) |
| *Durstenfeld, 2022, United States* | Chest pain, dyspnea, palpitations, or fatigue | 7 (18%) | 526 (464-553) | 23 | 21.2 ± 8.2<br>89 ± 23% | 16 | 28.8 ± 7.7<br>111 ± 20% |
| *Ladlow, 2022, United Kingdom* | Presence of 1 or more symptoms | 35 (31%) | 15 9 ± 7 | 61 | 32.4 ± 6.7 | 26 | 40.7 ± 8.9 |
| *Margalit, 2022, Israel<sup>44</sup>* | Fatigue | 14 (14%) | 240 ± 75 | 66 | 27.7 ± 7.5<br>96.1 ± 18.3% | 75 | 30.7 ± 7.5<br>99.6 ± 17.4% |
| *Skjørten, 2021, Norway<sup>42</sup>* | Dyspnea (mMRC 1-4) vs no dyspnea (mMRC=0) | 156 (100 %) | 104 (90-139) | 59 | 23.6 ± 7.9 | 67 | 31.9 ± 9.3 |
| *Szekely, 2021, Israel<sup>30</sup>* | Persistent symptoms (fatigue, dyspnea, or muscle weakness or pain) | NR<sup>a</sup> | 91 ± 26 | 8 | 1.5 ± 0.5 L/min | 23 | 1.7 ± 0.5 L/min |

Peak VO<sub>2</sub> is reported in ml/kg/min and percent predicted when available unless otherwise noted (Mohr, Szekely) as mean ± standard deviation or median (IQR). <sup>a</sup>All patients were evaluated in the emergency department but the number or percent admitted was not reported. Abbreviations: LC=Long COVID; mMRC=modified Medical Research Council Dyspnea Scale.

Nine studies which reported results for individuals with LC (n=464) and without (n=359) are summarized in Figure 2b. From meta-analyses of these studies, peak VO_2_ is estimated to be 4.9 ml/kg/min lower among individuals with versus without LC (95%CI -6.4 to -3.4). The I^2^ statistic (50%) suggests moderate heterogeneity. The heterogeneity is likely due to a combination of clinical variability and methodologic variability.

Clinical variability may result from the spectrum of LC severity and symptomatology.^37^ Although some studies suggest that acute severity may not impact exercise capacity,^38,39^ most studies suggest higher acuity is associated with worse exercise capacity.^31,36,40-42^ One study reported worse peak VO_2_ among unvaccinated compared to vaccinated individuals.^43^ Patients experience the total effect of SARS-CoV-2, but estimating the direct effect independent of hospitalization may be more helpful to understand pathophysiology. In addition, variability in definition of LC, time since infection, and enrollment criteria also resulted in heterogeneity, but lack of specificity in definition should bias results toward the null.

Age, sex, and BMI are highly associated with peak VO_2_ and likely associated with LC, but most studies did not adequately address confounding. Margalit reported that age, sex, pre-COVID-19 fitness levels and BMI were similar among those with and without LC and noted that post-infection exercise time per week was reduced by 2/3 among those with and without LC.^44^ Only two studies reported adjusted differences in peak VO_2_. Adjusted for gender, cardiovascular history, beta blockers, and aspirin, Barbagaleta estimated peak VO_2_ was 3.2 ml/min/kg lower in PASC (95% CI 0.9-5.5), but did not include BMI or age.^45^ Our own study reported that adjusted peak VO_2_ was 5.9 ml/kg/min (95% CI 2.3-9.6) lower in LC adjusted for age, sex, BMI, time since infection, and hospitalization. Because several studies did not report percent predicted peak VO_2_ we did not conduct our planned meta-analysis to address confounding.

### Patterns of Reduced Exercise Capacity

A common objective was to identify mechanisms of LC by characterizing patterns of CPET abnormalities. Nearly all studies defined “reduced exercise capacity” as <80% or 85% predicted, but few reported using specific guidelines or algorithms, or their classification approach. Because CPETs generate numerous data for each individual and there are different approaches to interpretation, notable differences in classification emerge between studies even with similar objective findings.

Deconditioning was the most commonly identified pattern and the main cause of reduced exercise capacity reported by 8 studies.^39,40,42,46-51^ Although ventilatory limitations were uncommon, multiple studies reported dysfunctional breathing or hyperventilation.^50,52-57^ Muscular/peripheral oxygen extraction abnormalities were also commonly reported.^26,29,58-62^ Using invasive CPET, Singh found reduced peripheral oxygen extraction, and others reported alterations in metabolism and lactate production.^27,58,60^ Cardiac limitations were uncommon, but Mancini reported attenuations in preload augmentation^52^ and Brown and Szekely reported reduced stroke volume augmentation.^30,33^ Five studies reported chronotropic incompetence to be a major cause of reduced exercise capacity,^25,30,44,63,64^ and most studies reported a lower peak heart rate among individuals with reduced exercise capacity. One study each specifically reported pulmonary fibrosis,^65^ pulmonary vascular limitation,^66^ impaired microcirculation,^61^ endothelial dysfunction,^67^ dysautonomia,^64^ and loss of mechanical efficiency^28^ as the primary etiology of reduced exercise capacity. Despite concerns about pulmonary thromboembolism during acute infection, pulmonary vascular limitations were uncommon.

### Longitudinal Trends

Three studies performed longitudinal CPETs in a subset; Cassar reported CPETs at 2-3 and 6 months; peak VO_2_ improved from 18.0 to 20.5 ml/kg/min but remained lower than controls (28.1 ml/kg/min, p≤0.001 for all).^29^ Evers found no significant changes in peak VO_2_ over 3 months among 23 individuals with reduced exercise capacity who underwent repeat CPET.^61^ Moulson found that peak VO_2_ improved among young symptomatic athletes 5 months after index study.^32^ Improvement correlated with symptom resolution, but peak heart rate was unexpectedly reduced. Barbara found a 15% improvement in peak VO_2_ after 8 weeks of cardiac rehabilitation.^23^

## Discussion

To this time, 39 studies performed CPET on over 2000 individuals after SARS-CoV-2 infection including nearly 1000 with LC and included 250 controls without SARS-CoV-2 infection. In meta-analysis of studies comparing infected individuals (mostly hospitalized, mostly with LC) to uninfected controls, we found a significantly lower peak VO_2_ and high heterogeneity, resulting in an estimate in which we have low confidence. In similar meta-analysis of LC versus recovered individuals, we found a modest but consistent effect suggesting that exercise capacity is reduced in LC. We identified themes in classification of exercise limitations without a single conclusive mechanism.

### Challenges to Estimating the Effect of SARS-CoV-2 on Exercise Capacity

Selection bias and lack of population-based sampling of infected persons bias estimates of the average causal effect of SARS-CoV-2 on exercise capacity. Included studies oversample hospitalized individuals with greater acute severity, more comorbidities, and lower baseline fitness. Hospitalization or need for intensive care is associated with greater reductions in peak VO_2_^31,36,40-42^ and with LC, but most with LC were not hospitalized.^68^ Differential selection bias may occur among those hospitalized, referred for clinical CPETs, or who attend CPET after joining a cohort. Selection bias likely results in overestimation of the proportion with reduced exercise capacity; only one study commented on the inability to estimate the proportion with reduced exercise capacity due to their sampling strategy.^45^

Most studies did not include contemporary uninfected controls. Ideally controls should be randomly sampled from the same target population as infected individuals, with selection criteria informed by the specific research question. Cohorts including non-hospitalized individuals may include uninfected individuals with similar demographics, comorbidities, and pre-infection fitness. To estimate effects independent of hospitalization, controls should include individuals hospitalized for similar conditions. Comparison with historical controls with known exercise capacity is particularly prone to bias effect estimates.

Few studies addressed confounding beyond implicitly adjusting for age, sex, height, and BMI by reporting percent predicted peak VO_2_. Likely confounders include age, sex, body mass index and composition, pre-infection fitness, and comorbid cardiac, pulmonary, and musculoskeletal conditions. None of the studies had pre-infection CPETs to compare within-individual change, which would provide stronger causal evidence. Margalit found no difference in recalled pre-infection weekly physical activity between those with and without LC.^44^ Two excluded studies among military recruits and professional athletes found reduced peak VO_2_ at 45-60 days post-infection compared to pre-infection.^69,70^ The main strategy to minimize confounding among studies with control groups is group matching on baseline characteristics (age, sex, weight, and comorbidities) or excluding those with pre-existing cardiopulmonary disease. A few studies used stepwise regression despite small sample sizes and co-linear variables, but no studies fully adjusted for likely confounders or reported adjusted differences in exercise capacity between those with and without COVID-19. Only two studies (including our own) presented an adjusted difference in peak VO_2_ between those with and without LC.^25,45^

Therefore, our confidence in the meta-analysis effect estimate of SARS-CoV-2 on exercise capacity is low. Our estimate likely exaggerates the true average causal effect, but the studies provide evidence that exercise capacity is reduced 3-6 months after hospitalization for COVID-19 and in LC compared to healthy controls. We have more confidence in our meta-analysis estimate of the difference in exercise capacity among those with and without LC. The included studies provide evidence of a clinically significant, mild-to-moderate decrease in exercise capacity among individuals with LC compared to infected individuals without LC despite different definitions of LC.

### Insights into Mechanisms of Reduced Exercise Capacity in LC

These studies should provide insight into mechanisms of LC, yet no consistent etiology of reduced exercise capacity has emerged, likely because of heterogeneity in inclusion criteria, variability in interpretation, and the likely presence of multiple mechanisms. Deconditioning, which occurs to some degree after any illness, was commonly identified. Identifying direct effects independent of debilitation is challenging. Using different exercise protocols or adjunctive measurements may result in differential classification. Submaximal exercise protocols make identifying chronotropic incompetence or peak VO_2_ challenging. Use of invasive CPET allows for estimation of cardiac output, preload, pulmonary hypertension, and peripheral oxygen extraction. Stress echocardiography or MRI approximates some of these measures. Some studies present findings according to mechanistic hypotheses (dysautonomia, endothelial function, dysfunctional breathing), all of which reported positive findings in accordance with their hypothesis.

Apart from deconditioning, commonly reported patterns include: (1) dysfunctional breathing or hyperventilation unexplained by baseline PFTs or findings on cross-sectional imaging, (2) changes in peripheral oxygen extraction/utilization, (3) chronotropic incompetence, and (4) lower stroke volume augmentation despite normal resting cardiac function. Apart from these, ventilatory, pulmonary vascular, and cardiac limitations are uncommon in LC, suggesting that direct heart or lung damage (especially given other negative testing) are not major drivers of exercise limitations. From the diversity of interpretations, different phenotypes resulting in exertional intolerance seem more likely than a single unifying mechanism.

Autonomic dysfunction and endothelial dysfunction are possible underlying mechanisms for these findings. One included study found endothelial dysfunction^67^ and two suggested dysautonomia^44,64^ are associated with reduced exercise capacity in LC. Dysfunctional breathing may also be a manifestation of dysautonomia.^64^ Peripheral vasomotor tone may be regulated by interaction between the autonomic system and endothelial function,^16^ so together they may explain differences in peripheral extraction. A small fiber neuropathy among individuals with LC-POTS may be associated with changes in cerebral blood flow and postural symptoms.^71,72^ Autonomic and endothelial dysfunction could be caused by SARS-CoV-2 infection of neurons and endothelial cells, chronic inflammation, or autoimmune mechanisms, all of which have been postulated in PASC, but no published studies include comprehensive autonomic testing, endothelial function, and CPET.

### Comparison with ME/CFS

Myalgic encephalitis/chronic fatigue syndrome (ME/CFS) is associated with reduced peak VO_2_, lower ventilatory efficiency, higher perceived exertion, and lower peak heart rates,^15^ and chronotropic incompetence contributes to exercise limitations.^14^ When matched by age and peak VO_2_, differences in heart rate are no longer statistically significant although adjusted heart rate reserve remained lower.^15^ However, if chronotropic incompetence reduces peak VO_2_, matching on peak VO_2_ would mask a true association. Alternatively, small fiber neuropathy causing peripheral shunting may cause reduced exercise capacity in ME/CFS.^16^ Whether LC has similar underlying pathophysiology as ME/CFS is unknown.

### Recommendations for CPET for LC Clinical Care and Research

CPET is clinically useful to narrow the differential diagnosis of exertional dyspnea, including in LC. A “normal” CPET without cardiopulmonary limitations will reassure some individuals with LC and increase comfort with physical activity. For those with objective limitations, identifying a cardiac or ventilatory limitation could provide clues for further diagnostic testing and treatment.

With regards to research, unanswered epidemiologic questions about the prevalence of exercise intolerance require intentional sampling to answer. Selection of control groups requires particular attention tailored to the research question. We recommend that CPET be performed as a maximal test that allows for assessment of chronotropy, except for those with severe post-exertional malaise, with adjunctive measures as per local expertise. Correlative data with autonomic testing may provide additional mechanistic insights. Given the high reproducibility within individuals and evidence of reduced exercise capacity in those with LC, CPET may be a useful measure to include in interventional trials for potential LC therapeutics.

### Limitations

We may have missed studies that met our inclusion criteria especially recent preprints. Many of the included studies were case-series, which contributed to classification of exercise limitations but not estimates of the prevalence or peak VO_2_. Because of selection bias, we could not estimate the prevalence of reduced exercise capacity. Although we conduced meta-analyses, the included studies likely overestimated the effect of SARS-CoV-2 infection. Finally, although we did not formally assess for publication bias given the small number of studies, only one study did not find a statistically significant effect; we cannot rule out publication bias contributing to exaggeration of effect estimates, although we mitigated this by including pre-prints and conference abstracts.

### Conclusions

In summary, we found evidence that exercise capacity is reduced after SARS-CoV-2 infection especially 3-6 months after hospitalization for severe acute COVID-19 and among those with LC. Further research should include longitudinal assessments to understand the trajectory of exercise capacity. Interventional trials of potential therapies are urgently needed including studies of rehabilitation to address deconditioning, as well as further mechanistic investigation into dysfunctional breathing, autonomic dysfunction, chronotropic incompetence, impaired oxygen uptake or utilization, and impaired stroke volume augmentation to identify treatments for LC.

## Supporting information

PRISMA Checklist

## Data Availability

All data produced in the present study are available upon reasonable request to the authors.

## Supplemental Methods

### Protocol

The full, pre-registered Protocol is available at https://www.crd.york.ac.uk/prospero/display_record.php?ID=CRD42021299842.

This review was conducted according to the PRISMA (Preferred Reporting Items for Systematic Reviews and Meta-Analysis)^22^ guidelines and was registered prospectively on PROSPERO prior to beginning the search.

#### Condition being studied

post-acute sequelae of COVID-19, also known as Long COVID, which according to the WHO definition is >3 months after acute infection with SARS-CoV-2.

#### Inclusion criteria

all studies of adults with confirmed COVID-19 at least 3 months after onset that include cardiopulmonary exercise testing with measurement of peak VO_2_ published since 2020 will be included. Baseline cardiopulmonary exercise testing from interventional or randomized controlled trials will also be included if they meet the other inclusion criteria.

#### Exclusion criteria

studies of children, studies of conditions other than COVID-19/SARS-CoV-2, studies in the acute or early post-acute phase (<3 months after infection), review articles, case reports.

#### Intervention/exposure

Cardiopulmonary exercise testing, which includes measurement of metabolic gases with either treadmill or cycle ergometer exercise.

#### Participants/population

We are interested in all adults with COVID without respect to hospitalization status or severity of acute illness.

#### Inclusion/Exclusion criteria

adults with confirmed COVID-19 at least 3 months after onset that include cardiopulmonary exercise testing with measurement of peak VO_2_ will be included. We excluded studies of children, studies of conditions other than COVID-19/SARS-CoV-2, studies in the acute phase (<3 months after infection).

#### Comparators/control

We will include case series without controls, as well as studies with healthy controls, control participants with unexplained dyspnea, or that compare those who have fully recovered from COVID compared to those reporting ongoing symptoms.

#### Types of studies to be included

We will include observational studies including case series, cross-sectional studies, case-control studies, and cohort studies. We will also include randomized trials of interventions, in which case we will use baseline CPET data. We will exclude case reports and review articles.

#### Context

We will include studies that include the full spectrum of COVID-19; specifically, we will not restrict to only studies of those requiring ICU or hospitalization during acute infection.

#### Main Outcomes

The primary outcome will be peak VO_2_ (in ml/kg/min and % predicted). If meta-analysis is possible, studies that do not include this measure will be excluded from meta-analysis. We will report the difference in peak VO_2_ between those with and without COVID and among those with COVID between those with and without post-acute sequelae.

#### Additional outcomes

Additional outcomes will include the proportion with exercise limitation <80 or 85% of predicted (different studies use different cutoffs), difference in exercise capacity between those with and without cardiopulmonary symptoms (absolute and relative difference with 95% confidence intervals and p value), common features among those with limitations (i.e., reduced oxygen pulse pressure, reduced chronotropic response). We will likely report these effect measures in odds-ratios as we expect that many of the studies may be case-control studies.

#### Search Strategy & Information Sources

A comprehensive, electronic search strategy will be used to identify studies published since 2020 and indexed in PubMed, EMBASE, and Web of Science by a research librarian (PT) with extensive experience in systematic reviews. Unpublished abstracts from conference proceedings and indexed preprints will be included as part of our gray literature search. We will also review references from studies selected for data extraction. The search strategy will include terms and synonyms for the following: (COVID or SARS-CoV-2) AND (“cardiopulmonary exercise test*” OR (CPET or CPX or CPEX) OR exercise capacity OR VO2 OR anaerobic threshold). Searches will be tailored to each database depending on indexing terminology. Searches were conducted on December 20, 2021, and rerun prior to the final analysis on May 24, 2022; pre-prints were searched through June 9, 2022. Abstracts were reviewed for inclusion by two independent reviewers (MSD & KS); if there is disagreement after consensus discussion, a third reviewer will be consulted. All data extraction was done independently, in duplicate, using REDCap for data entry.

#### Gray literature plan

see search strategy for details; we will review conference abstracts, pre-prints, and references from studies that meet the inclusion criteria.

#### Data Extraction (Selection & Coding)

Data including authors, title, date of study, location of study, sample size (including total with COVID, total with Cardiopulmonary Long COVID, and COVID-negative controls, if included), median time since acute infection and interquartile range, inclusion criteria (with particular attention to inclusion of hospitalized/ICU/ambulatory during acute illness and those with specific comorbidities or populations of interest), comparator group, exercise modality (treadmill or cycle ergometer), peak VO2 (in ml/kg/min and % predicted), proportion with exercise limitation <85% of predicted, difference in exercise capacity between those with and without cardiopulmonary symptoms (absolute and relative difference with 95% confidence intervals and p value), common features among those with limitations (i.e., reduced oxygen pulse pressure, reduced chronotropic response). If available, other cardiopulmonary parameters will be recorded including echocardiographic, pulmonary function tests, chest computed tomography, and cardiac magnetic resonance imaging.

#### Data Management

Studies identified through the searches will be managed using Covidence. Data extracted will be recorded using REDCap.

#### Quality Assessment

We will use Cochrane’s Quality in Prognostic Studies (QUIPS) tool to assess for bias of included studies. We will assess study populations (especially choice of control groups), study attrition for non-cross-sectional studies, peak VO_2_ assessment quality, outcome measurement, study confounding, and statistical analysis and reporting. We will use Cochrane’s Quality in Prognostic Studies (QUIPS) tool to assess for bias of included studies.

#### Data synthesis

Overall findings of each study will be summarized in a table. If possible, a meta-analysis will be performed to compare the peak VO2 among those with and without COVID. An odds ratio of having reduced exercise capacity may also be estimated if possible. Heterogeneity will be assessed using I^2^. The primary subgroup we plan to investigate is to compare peak VO2 (and the other explanatory variables for reduced exercise capacity) among those with and without PASC/Long COVID. If possible, we may also compare those with severe acute infection requiring hospitalization and/or ICU care with those who were asymptomatic or had mild acute infection. Lastly, we may compare the early post-acute period (3-6 months), medium term (6-12 months), and long term (>12 months). Analyses will be performed using STATA version 17.

#### Analysis of subgroups

The primary subgroup we plan to investigate is to compare peak VO2 (and the other explanatory variables for reduced exercise capacity) among those with and without PASC/Long COVID. If possible, we may also compare those with severe acute infection requiring hospitalization and/or ICU care with those who were asymptomatic or had mild acute infection. Lastly, we may compare the early post-acute period (3-6 months), medium term (6-12 months), and long term (>12 months).

#### Risk of Bias/Quality Assessment

Risk of bias will be assessed at both the study and the outcome level for each included study. Publication bias will be assessed using a Funnel Plot. The strength of the body of evidence will be assessed using the GRADE (Grading of Recommendations, Assessment, Development, and Evaluations) framework.

**Supplemental Table 1:** Search Appendix

| DATABASE | SEARCH STRATEGY |
| --- | --- |
| PubMed | ("COVID-19"[Mesh] OR COVID OR "SARS-CoV-2"[Mesh] OR SARS-CoV-2) AND ("cardiopulmonary exercise test*" OR CPET OR CPX OR CPEX OR exercise capacity OR VO2 OR "Anaerobic Threshold"[Mesh] OR anaerobic threshold) |
| Web of Science | (COVID OR SARS-CoV-2) AND ("cardiopulmonary exercise test*" OR CPET OR CPX OR CPEX OR exercise capacity OR VO2 OR anaerobic threshold) |
| Embase | ('coronavirus disease 2019'/exp OR 'coronavirus disease 2019') AND ('cardiopulmonary exercise test'/exp OR 'cardiopulmonary exercise test' OR 'cardiopulmonary exercise testing'/exp OR 'cardiopulmonary exercise testing' OR cpet OR cpx OR cpex OR 'exercise capacity'/exp OR 'exercise capacity' OR vo2 OR 'anaerobic threshold'/exp OR 'anaerobic threshold') |

**Supplemental Table 2:**
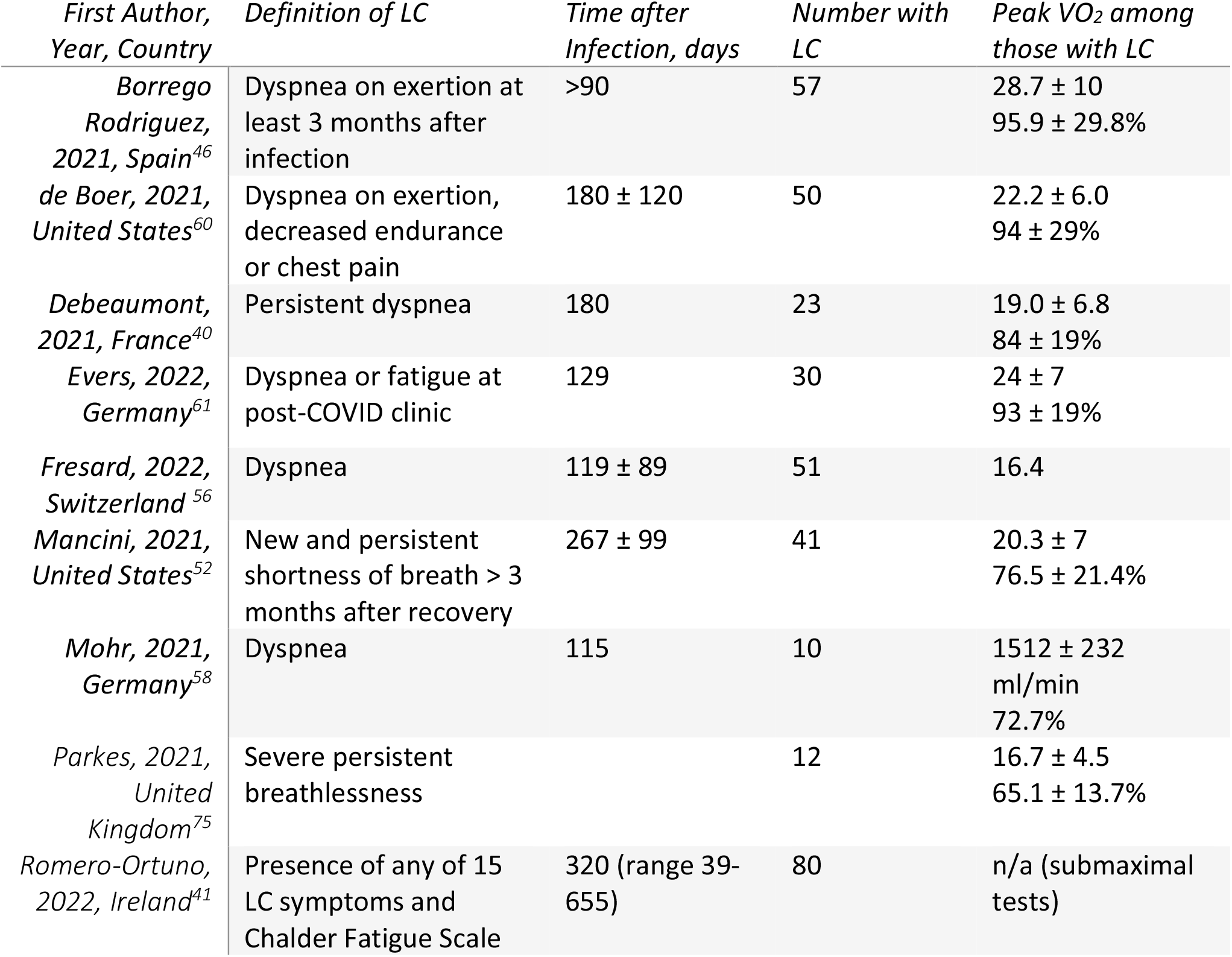
Case series of individuals with LC

| *First Author,*<br>*Year, Country* | *Definition of LC* | *Time after*<br>*Infection, days* | *Number with*<br>*LC* | *Peak VO<sub>2</sub> among*<br>*those with LC* |
| --- | --- | --- | --- | --- |
| *Borrego*<br>*Rodriguez,*<br>*2021, Spain<sup>46</sup>* | Dyspnea on exertion at<br>least 3 months after<br>infection | >90 | 57 | 28.7 ± 10<br>95.9 ± 29.8% |
| *de Boer, 2021,*<br>*United States<sup>60</sup>* | Dyspnea on exertion,<br>decreased endurance<br>or chest pain | 180 ± 120 | 50 | 22.2 ± 6.0<br>94 ± 29% |
| *Debeaumont,*<br>*2021, France<sup>40</sup>* | Persistent dyspnea | 180 | 23 | 19.0 ± 6.8<br>84 ± 19% |
| *Evers, 2022,*<br>*Germany<sup>61</sup>* | Dyspnea or fatigue at<br>post-COVID clinic | 129 | 30 | 24 ± 7<br>93 ± 19% |
| *Fresard, 2022,*<br>*Switzerland<sup>56</sup>* | Dyspnea | 119 ± 89 | 51 | 16.4 |
| *Mancini, 2021,*<br>*United States<sup>52</sup>* | New and persistent<br>shortness of breath > 3<br>months after recovery | 267 ± 99 | 41 | 20.3 ± 7<br>76.5 ± 21.4% |
| *Mohr, 2021,*<br>*Germany<sup>58</sup>* | Dyspnea | 115 | 10 | 1512 ± 232<br>ml/min<br>72.7% |
| *Parkes, 2021,*<br>*United*<br>*Kingdom<sup>75</sup>* | Severe persistent<br>breathlessness |  | 12 | 16.7 ± 4.5<br>65.1 ± 13.7% |
| *Romero-Ortuno,*<br>*2022, Ireland<sup>41</sup>* | Presence of any of 15<br>LC symptoms and<br>Chalder Fatigue Scale | 320 (range 39-655) | 80 | n/a (submaximal<br>tests) |

## References

1. Groff D, Sun A, Ssentongo AE, et al. Short-term and Long-term Rates of Postacute Sequelae of SARS-CoV-2 Infection: A Systematic Review. JAMA Netw Open. 2021;4(10):e2128568.

2. Hirschtick JL, Titus AR, Slocum E, et al. Population-Based Estimates of Post-acute Sequelae of Severe Acute Respiratory Syndrome Coronavirus 2 (SARS-CoV-2) Infection (PASC) Prevalence and Characteristics. Clin Infect Dis. 2021;73(11):2055–2064.

3. Taquet M, Dercon Q, Luciano S, Geddes JR, Husain M, Harrison PJ. Incidence, co-occurrence, and evolution of long-COVID features: A 6-month retrospective cohort study of 273,618 survivors of COVID-19. PLoS Med. 2021;18(9):e1003773.

4. Piotr DA, Gaughan PC. Technical article: Updated estimates of the prevalence of post-acute symptoms among people with coronavirus (COVID-19) in the UK: 26 April 2020 to 1 August 2021. In: Statistics UOfN, ed. London2021.

5. Yomogida K, Zhu S, Rubino F, Figueroa W, Balanji N, Holman E. Post-Acute Sequelae of SARS-CoV-2 Infection Among Adults Aged >/=18 Years - Long Beach, California, April 1-December 10, 2020. MMWR Morb Mortal Wkly Rep. 2021;70(37):1274–1277.

6. Xie Y, Bowe B, Al-Aly Z. Burdens of post-acute sequelae of COVID-19 by severity of acute infection, demographics and health status. Nat Commun. 2021;12(1):6571.

7. Al-Aly Z, Bowe B, Xie Y. Long COVID after breakthrough SARS-CoV-2 infection. Nature Medicine. 2022.

8. Huang L, Yao Q, Gu X, et al. 1-year outcomes in hospital survivors with COVID-19: a longitudinal cohort study. The Lancet. 2021;398(10302):747–758.

9. American Thoracic S, American College of Chest P. ATS/ACCP Statement on cardiopulmonary exercise testing. Am J Respir Crit Care Med. 2003;167(2):211–277.

10. Wasserman K HJ, Sue DY, Stringer W, Whipp BJ. Principles of Exercise Testing and Interpretation. 4th ed. Philadelpha: Lippincott Williams and Wilkins; 2005.

11. Balady GJ, Arena R, Sietsema K, et al. Clinician’s Guide to cardiopulmonary exercise testing in adults: a scientific statement from the American Heart Association. Circulation. 2010;122(2):191–225.

12. Chan VL, Lam JY, Leung W-S, Lin AW, Chu C-M. Exercise Limitation in Survivors of Severe Acute Respiratory Syndrome (SARS). Chest. 2004;126(4).

13. Patterson AJ, Sarode A, Al-Kindi S, et al. Evaluation of dyspnea of unknown etiology in HIV patients with cardiopulmonary exercise testing and cardiovascular magnetic resonance imaging. J Cardiovasc Magn Reson. 2020;22(1):74.

14. Davenport TE, Lehnen M, Stevens SR, VanNess JM, Stevens J, Snell CR. Chronotropic Intolerance: An Overlooked Determinant of Symptoms and Activity Limitation in Myalgic Encephalomyelitis/Chronic Fatigue Syndrome? Front Pediatr. 2019;7:82.

15. Cook DB, VanRiper S, Dougherty RJ, et al. Cardiopulmonary, metabolic, and perceptual responses during exercise in Myalgic Encephalomyelitis/Chronic Fatigue Syndrome (ME/CFS): A Multi-site Clinical Assessment of ME/CFS (MCAM) sub-study. PLOS ONE. 2022;17(3):e0265315.

16. Joseph P, Arevalo C, Oliveira RKF, et al. Insights From Invasive Cardiopulmonary Exercise Testing of Patients With Myalgic Encephalomyelitis/Chronic Fatigue Syndrome. Chest. 2021;160(2):642–651.

17. Malhotra R, Bakken K, D’Elia E, Lewis GD. Cardiopulmonary Exercise Testing in Heart Failure. JACC Heart Fail. 2016;4(8):607–616.

18. Levett DZH, Jack S, Swart M, et al. Perioperative cardiopulmonary exercise testing (CPET): consensus clinical guidelines on indications, organization, conduct, and physiological interpretation. Br J Anaesth. 2018;120(3):484–500.

19. Baratto C, Caravita S, Faini A, et al. Impact of COVID-19 on exercise pathophysiology: a combined cardiopulmonary and echocardiographic exercise study. J Appl Physiol (1985). 2021;130(5):1470–1478.

20. Arena R, Faghy MA. Cardiopulmonary exercise testing as a vital sign in patients recovering from COVID-19. Expert Review of Cardiovascular Therapy. 2021;19(10):877–880.

21. Soriano JM, J; Diaz JV; Murthy, S;; Relan, P. A clinical case definition of post COVID-19 condition by a Delphi consensus. In: Organization WH, ed. 2021. 1 ed: ; 2021.

22. Shamseer L, Moher D, Clarke M, et al. Preferred reporting items for systematic review and meta-analysis protocols (PRISMA-P) 2015: elaboration and explanation. BMJ. 2015;350:g7647.

23. Barbara C, Clavario P, De Marzo V, et al. Effects of exercise rehabilitation in patients with long COVID-19. Eur J Prev Cardiol. 2022;29(7):e258–e260.

24. Clavario P, De Marzo V, Lotti R, et al. Cardiopulmonary exercise testing in COVID-19 patients at 3 months follow-up. Int J Cardiol. 2021;340:113–118.

25. Durstenfeld MS, Peluso MJ, Kaveti P, et al. Inflammation during early post-acute COVID-19 is associated with reduced exercise capacity and Long COVID symptoms after 1 year. Preprint at medrxivorg https://doiorg/101101/2022051722275235 (Posted May 19, 2022). 2022.

26. Alba GA, Ziehr DR, Rouvina JN, et al. Exercise performance in patients with post-acute sequelae of SARS-CoV-2 infection compared to patients with unexplained dyspnea. EClinicalMedicine. 2021;39:101066.

27. Singh I, Joseph P, Heerdt PM, et al. Persistent Exertional Intolerance After COVID-19: Insights From Invasive Cardiopulmonary Exercise Testing. Chest. 2022;161(1):54–63.

28. Pleguezuelos E, Del Carmen A, Llorensi G, et al. Severe loss of mechanical efficiency in COVID-19 patients. Journal of Cachexia, Sarcopenia and Muscle. 2021;12(4):1056–1063.

29. Cassar MP, Tunnicliffe EM, Petousi N, et al. Symptom Persistence Despite Improvement in Cardiopulmonary Health - Insights from longitudinal CMR, CPET and lung function testing post-COVID-19. EClinicalMedicine. 2021;41:101159.

30. Szekely Y, Lichter Y, Sadon S, et al. Cardiorespiratory Abnormalities in Patients Recovering from Coronavirus Disease 2019. J Am Soc Echocardiogr. 2021;34(12):1273-1284.e1279.

31. Ladlow P, O’Sullivan O, Bennett AN, et al. The effect of medium-term recovery status after COVID-19 illness on cardiopulmonary exercise capacity in a physically active adult population. J Appl Physiol (1985). 2022.

32. Moulson N, Gustus SK, Scirica C, et al. Diagnostic evaluation and cardiopulmonary exercise test findings in young athletes with persistent symptoms following COVID-19. Br J Sports Med. 2022.

33. Brown JT, Saigal A, Karia N, et al. Ongoing Exercise Intolerance Following COVID-19: A Magnetic Resonance-Augmented Cardiopulmonary Exercise Test Study. J Am Heart Assoc. 2022;11(9):e024207.

34. Cassar MP, Lewandowski AJ, Mahmod M, et al. Longitudinal trajectory of cardiac magnetic resonance and cardiopulmonary exercise testing findings in moderate to severe COVID-19 and association with symptoms. European Heart Journal. 2021;42(Supplement_1).

35. Raman B, Cassar MP, Tunnicliffe EM, et al. Medium-term effects of SARS-CoV-2 infection on multiple vital organs, exercise capacity, cognition, quality of life and mental health, post-hospital discharge. EClinicalMedicine. 2021;31:100683.

36. Vonbank K, Lehmann A, Bernitzky D, et al. Predictors of Prolonged Cardiopulmonary Exercise Impairment After COVID-19 Infection: A Prospective Observational Study. Front Med (Lausanne). 2021;8:773788.

37. Peluso MJ, Kelly JD, Lu S, et al. Persistence, Magnitude, and Patterns of Postacute Symptoms and Quality of Life Following Onset of SARS-CoV-2 Infection: Cohort Description and Approaches for Measurement. Open Forum Infect Dis. 2022;9(2):ofab640.

38. Rinaldo RF, Mondoni M, Parazzini EM, et al. Severity does not impact on exercise capacity in COVID-19 survivors. Respir Med. 2021;187:106577.

39. Ribeiro Baptista B, d’Humieres T, Schlemmer F, et al. Identification of factors impairing exercise capacity after severe COVID-19 pulmonary infection: a 3-month follow-up of prospective COVulnerability cohort. Respir Res. 2022;23(1):68.

40. Debeaumont D, Boujibar F, Ferrand-Devouge E, et al. Cardiopulmonary Exercise Testing to Assess Persistent Symptoms at 6 Months in People With COVID-19 Who Survived Hospitalization: A Pilot Study. Phys Ther. 2021;101(6).

41. Romero-Ortuno R, Jennings G, Xue F, Duggan E, Gormley J, Monaghan A. Predictors of Submaximal Exercise Test Attainment in Adults Reporting Long COVID Symptoms. J Clin Med. 2022;11(9).

42. Skjorten I, Ankerstjerne OAW, Trebinjac D, et al. Cardiopulmonary exercise capacity and limitations 3 months after COVID-19 hospitalisation. Eur Respir J. 2021;58(2).

43. Blumberg Y, Edelstein M, Jabal KA, et al. Protective effect of BNT162b2 vaccination on aerobic capacity following mild to moderate SARS-CoV-2 infection: a cross sectional study, Israel, March-December 2021. medRxiv. 2022:2021.2012.2030.21268538.

44. Margalit I, Yelin D, Sagi M, et al. Risk factors and multidimensional assessment of long COVID fatigue: a nested case-control study. Clin Infect Dis. 2022.

45. Barbagelata L, Masson W, Iglesias D, et al. Cardiopulmonary Exercise Testing in Patients with Post-COVID-19 Syndrome. Med Clin (Barc). 2021.

46. Borrego Rodriguez J, Berenguel Senen A, De Cabo Porras C, et al. Cardiopulmonary exercise test in patients with persistent dyspnea after COVID-19 disease. European Heart Journal. 2021;42(Supplement_1).

47. Godinho L, Freeman A. S25 Cardiopulmonary exercise testing to evaluate exercise limitation and shortness of breath in long COVID. Thorax. 2021;76(Suppl 2):A19–A20.

48. Jahn K, Sava M, Sommer G, et al. Exercise capacity impairment after COVID-19 pneumonia is mainly caused by deconditioning. Eur Respir J. 2022;59(1):2101136.

49. Johnsen S, Sattler SM, Miskowiak KW, et al. Descriptive analysis of long COVID sequelae identified in a multidisciplinary clinic serving hospitalised and non-hospitalised patients. ERJ Open Res. 2021;7(3):00205–02021.

50. Motiejunaite J, Balagny P, Arnoult F, et al. Hyperventilation as one of the mechanisms of persistent dyspnoea in SARS-CoV-2 survivors. Eur Respir J. 2021;58(2):2101578.

51. Rinaldo RF, Mondoni M, Parazzini EM, et al. Deconditioning as main mechanism of impaired exercise response in COVID-19 survivors. Eur Respir J. 2021;58(2):2100870.

52. Mancini DM, Brunjes DL, Lala A, Trivieri MG, Contreras JP, Natelson BH. Use of Cardiopulmonary Stress Testing for Patients With Unexplained Dyspnea Post-Coronavirus Disease. JACC Heart Fail. 2021;9(12):927–937.

53. Aparisi A, Ybarra-Falcon C, Garcia-Gomez M, et al. Exercise Ventilatory Inefficiency in Post-COVID-19 Syndrome: Insights from a Prospective Evaluation. J Clin Med. 2021;10(12).

54. Crisafulli E, Dorelli G, Sartori G, Dalle Carbonare L. Exercise ventilatory inefficiency may be a relevant CPET-feature in COVID-19 survivors. Int J Cardiol. 2021;343:200.

55. Dorelli G, Braggio M, Gabbiani D, et al. Importance of Cardiopulmonary Exercise Testing amongst Subjects Recovering from COVID-19. Diagnostics (Basel). 2021;11(3).

56. Fresard I, Genecand L, Altarelli M, et al. Dysfunctional breathing diagnosed by cardiopulmonary exercise testing in ‘long COVID’ patients with persistent dyspnoea. BMJ Open Respir Res. 2022;9(1):e001126.

57. von Gruenewaldt A, Nylander E, Hedman K. Classification and occurrence of an abnormal breathing pattern during cardiopulmonary exercise testing in subjects with persistent symptoms following COVID-19 disease. Physiol Rep. 2022;10(4):e15197.

58. Mohr A, Dannerbeck L, Lange TJ, et al. Cardiopulmonary exercise pattern in patients with persistent dyspnoea after recovery from COVID-19. Multidiscip Respir Med. 2021;16(1):732.

59. Clavario P, De Marzo V, Lotti R, et al. Cardiopulmonary exercise testing in COVID-19 patients at 3 months follow-up. Int J Cardiol. 2021;340:113–118.

60. de Boer E, Petrache I, Goldstein NM, et al. Decreased Fatty Acid Oxidation and Altered Lactate Production during Exercise in Patients with Post-acute COVID-19 Syndrome. Am J Respir Crit Care Med. 2022;205(1):126–129.

61. Evers G, Schulze AB, Osiaevi I, et al. Sustained Impairment in Cardiopulmonary Exercise Capacity Testing in Patients after COVID-19: A Single Center Experience. Can Respir J. 2022;2022:2466789.

62. Vannini L, Quijada-Fumero A, Martín MPR, Pina NC, Afonso JSH. Cardiopulmonary exercise test with stress echocardiography in COVID-19 survivors at 6 months follow-up. Eur J Intern Med. 2021;94:101–104.

63. Abdallah SJ, Voduc N, Corrales-Medina VF, et al. Symptoms, Pulmonary Function, and Functional Capacity Four Months after COVID-19. Ann Am Thorac Soc. 2021;18(11):1912–1917.

64. Ladlow P, O’Sullivan O, Houston A, et al. Dysautonomia following COVID-19 is not associated with subjective limitations or symptoms but is associated with objective functional limitations. Heart Rhythm. 2022;19(4):613–620.

65. Liu M, Lv F, Huang Y, Xiao K. Follow-Up Study of the Chest CT Characteristics of COVID-19 Survivors Seven Months After Recovery. Front Med (Lausanne). 2021;8:636298.

66. Kersten J, Baumhardt M, Hartveg P, et al. Long COVID: Distinction between Organ Damage and Deconditioning. Journal of Clinical Medicine. 2021;10(17):3782.

67. Ambrosino P, Parrella P, Formisano R, et al. Cardiopulmonary Exercise Performance and Endothelial Function in Convalescent COVID-19 Patients. J Clin Med. 2022;11(5).

68. Sudre CH, Murray B, Varsavsky T, et al. Attributes and predictors of long COVID. Nat Med. 2021;27(4):626–631.

69. Crameri GAG, Bielecki M, Zust R, Buehrer TW, Stanga Z, Deuel JW. Reduced maximal aerobic capacity after COVID-19 in young adult recruits, Switzerland, May 2020. Euro Surveill. 2020;25(36):2001542.

70. Parpa K, Michaelides M. The Effect of COVID-19 Infection on the Aerobic Capacity of Professional Soccer Players. Preprint at https://doiorg/1021203/rs3rs-1034685/v1 (Posted November 15th, 2021). 2021.

71. Novak P, Mukerji SS, Alabsi HS, et al. Multisystem Involvement in Post-Acute Sequelae of Coronavirus Disease 19. Ann Neurol. 2022;91(3):367–379.

72. Oaklander AL, Mills AJ, Kelley M, et al. Peripheral Neuropathy Evaluations of Patients With Prolonged Long COVID. Neurol Neuroimmunol Neuroinflamm. 2022;9(3).

73. Schaeffer MR, Cowan J, Milne KM, et al. Cardiorespiratory physiology, exertional symptoms, and psychological burden in post-COVID-19 fatigue. Respir Physiol Neurobiol. 2022;302:103898.

74. Blokland IJ, Ilbrink S, Houdijk H, et al. [Exercise capacity after mechanical ventilation because of COVID-19: Cardiopulmonary exercise tests in clinical rehabilitation]. Ned Tijdschr Geneeskd. 2020;164.

75. Parkes E, Shakespeare J, Robbins T, Kyrou I, Randeva H, Ali A. Utility of Cardiopulmonary Exercise Testing (CPET) in the Post-COVID-19 Context: Retrospective Analysis of a Single Centre Experience. Preprint at https://doiorg/1021203/rs3rs-537361/v1 (Posted May 25th, 2021). 2021.

